# SARS-CoV-2 R.1 lineage variants prevailed in Tokyo in March 2021

**DOI:** 10.1101/2021.05.11.21257004

**Authors:** Katsutoshi Nagano, Chihiro Tani-Sassa, Yumi Iwasaki, Yuna Takatsuki, Sonoka Yuasa, Yuta Takahashi, Jun Nakajima, Kazunari Sonobe, Naoya Ichimura, Yoko Nukui, Hiroaki Takeuchi, Kousuke Tanimoto, Yukie Tanaka, Akinori Kimura, Shuji Tohda

## Abstract

**Background:** The spread of SARS-CoV-2 variants, such as B.1.1.7 and B.1.351, has become a crucial issue worldwide. Therefore, we began testing all patients with COVID-19 for the N501Y and E484K mutations by using polymerase chain reaction (PCR)-based methods.

**Methods:** Nasopharyngeal swab samples from 108 patients who visited our hospital between February and April 2021 were analyzed. The samples were analyzed using reverse transcription-PCR with melting curve analysis to detect the N501Y and E484K mutations. A part of the samples were also subjected to whole genome sequencing (WGS). Clinical parameters such as mortality and admission to the intensive care unit were analyzed to examine the association between increased disease severity and the E484K mutation.

**Results:** The ratio of cases showing the 501N+484K mutation rapidly increased from 8% in February to 46% in March. WGS revealed that the viruses with 501N+484K mutation are R.1 lineage variants. Evidence of increased disease severity related to the R.1 variants were not found.

**Conclusions:** We found that the R.1 lineage variants rapidly prevailed in Tokyo in March 2021, which suggests the increased transmissibility of R.1 variants while they showed no increased severity.

## 1. introduction

Since the beginning of 2021, the spread of severe acute respiratory syndrome coronavirus 2 (SARS-CoV-2) variants such as B.1.1.7 known as United Kingdom (UK) type, B.1.351 (South Africa type), and P.1 (Brazil type) has become a crucial issue worldwide.^1-4^ We are also aware of this issue at our hospital, Tokyo Medical and Dental University Hospital, which mainly treats patients with severe coronavirus disease 2019 (COVID-19) from all over Tokyo.

Therefore, we began testing all samples from outpatients and inpatients who tested polymerase chain reaction (PCR)-positive for SARS-CoV-2 for its associated mutations, N501Y and E484K, using PCR-based melting curve analysis. We found that the conventional strains (European lineage) with the 501N+484E which accounted for the majority of cases until February were replaced by R.1 variants with 501N+484K mutation in March. Since studies of the R.1 variants have rarely been reported, here we present the findings from patients with COVID-19 treated at our hospital.

## 2. Material and methods

This study included 108 patients with COVID-19 who visited our hospital from February to April 2021. The nasopharyngeal swab samples obtained from patients were immersed in test tubes with 1 mL of phosphate-buffered saline containing 1% dithiothreitol. To detect SARS-CoV-2 in the samples, one-step reverse transcription-quantitative PCR (RT-qPCR) was performed without viral RNA purification using the 2019 Novel Coronavirus Detection Kit (Shimadzu Corp.; Kyoto, Japan) and the QuantStudio 5 Dx Real-Time PCR System (Thermo Fisher Scientific, Inc.; Waltham, MA, USA).

To detect N501Y and E484K mutations, viral RNA was purified from the PCR-positive samples using the EZ1 Virus Mini Kit v2.0 and EZ1 advanced XL (QIAGEN; Venlo, Netherlands). RT-PCR was performed using the primers and probes provided in the VirSNiP SARS-CoV-2 Spike N501Y and VirSNiP SARS-CoV-2 Spike E484K kits (TIB Molbiol; Berlin, Germany) and the LightCycler Multiplex RNA Virus Master (Roche Molecular Systems, Inc.; Basel, Switzerland). The Melting curve analyses of the PCR products were performed according to the manufacturer’s instructions, and the 501N, 501Y, 484E, and 484K types were determined by the melting temperature.

Using a part of the samples that showed 501N+484E, 501N+484K, and 501Y+484E, whole genome sequencing (WGS) was performed using a next-generation sequencer to specify the lineages.

To investigate whether the 501N+484K variants are involved in the increased severity of the disease, we compared elements from the patients’ clinical profiles, such as mortality and admission to the intensive care unit (ICU), between 501N+484E and 501N+484K patients who were discharged by April 30.

This study was approved by Medical Research Ethics Committee of Tokyo Medical and Dental University (approval number: M2020-004) and was conducted in accordance with the ethical standards of the 1964 Helsinki declaration.

## 3. Results

The number of cases by the three variant types in each month is shown in Figure 1(A). Samples for which the 501 or 484 type could not be determined were classified as untyped. In February, the 501N+484E type accounted for 80% of cases and the 501N+484K type for 8%. In March, the 501N+484K type increased to 46% and the 501Y+484E type appeared at the end of March. In April, along with the rapid increase of patients with COVID-19, the 501Y+484E variant type increased and accounted for about half of all cases. The type of 501Y+484K, which corresponds to the B.1.351 and P.1 variants, were not found during the three-month study period.

**FIGURE 1.**
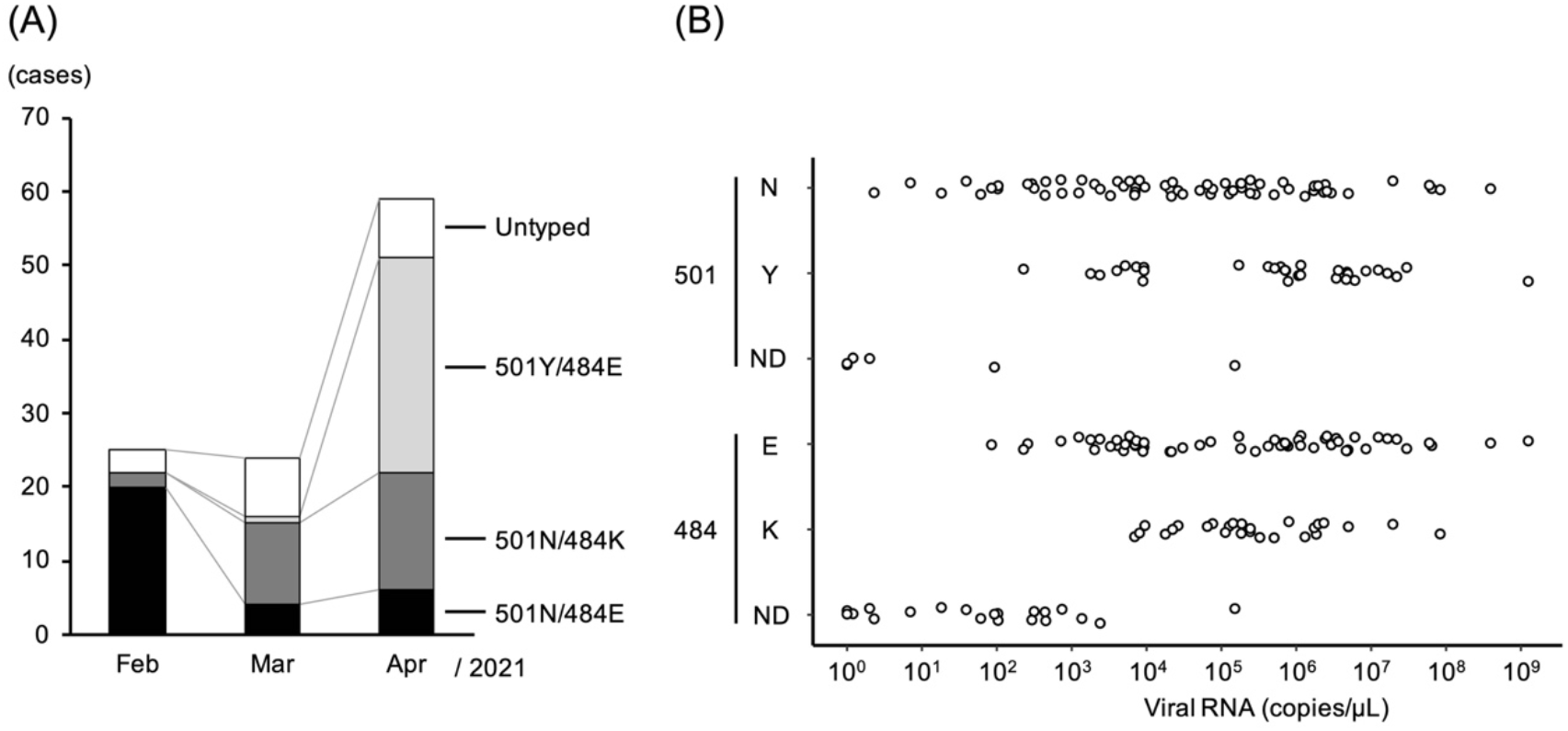
Number of cases by each variant type determined by PCR-based melting curve analysis (A) and copy numbers of viral RNA in swab-soaked samples showing 501N, 501Y, 484E, 484K types and samples of which types could not be determined (B). ND, not determined.

Figure 1(B) shows the distribution of the copy numbers from the samples showing 501N, 501Y, 484E, and 484K and the untyped samples that did not produce enough PCR products for melting curve analysis. There were no significant differences in the copy numbers between the 501N and 501Y samples or between the 484E and 484K samples. This figure also suggests that the 501 and 484 types may not be determined if the viral RNA concentration in swab-soaked samples is approximately less than 100 copies/μL or less than 1,000 copies/μL, respectively, by our method.

WGS analysis of the selected samples revealed that the 501N+484K type is an R.1 lineage (W152L, E484K, D614G, G769V) variant, which is the sublineage of B.1.1.316. The 501Y+484E variant type is from the B.1.1.7 lineage (UK lineage) and the 501N+484E type is from the B.1.1.214 lineage (Japan lineage).

Table 1 shows the clinical profiles of 501N+484E (Japan lineage) and 501N+484K (R.1 lineage) cases. There were no significant differences in any of the variables between the two groups. The length of hospitalization tended to be short in the 501N+484K group, since this group included four patients who did not need hospitalization. The profiles of 501Y+484E (UK lineage) cases were not listed because approximately half of the patients were still in the hospital and their outcome had not been determined at the end of this study.

**TABLE 1.**
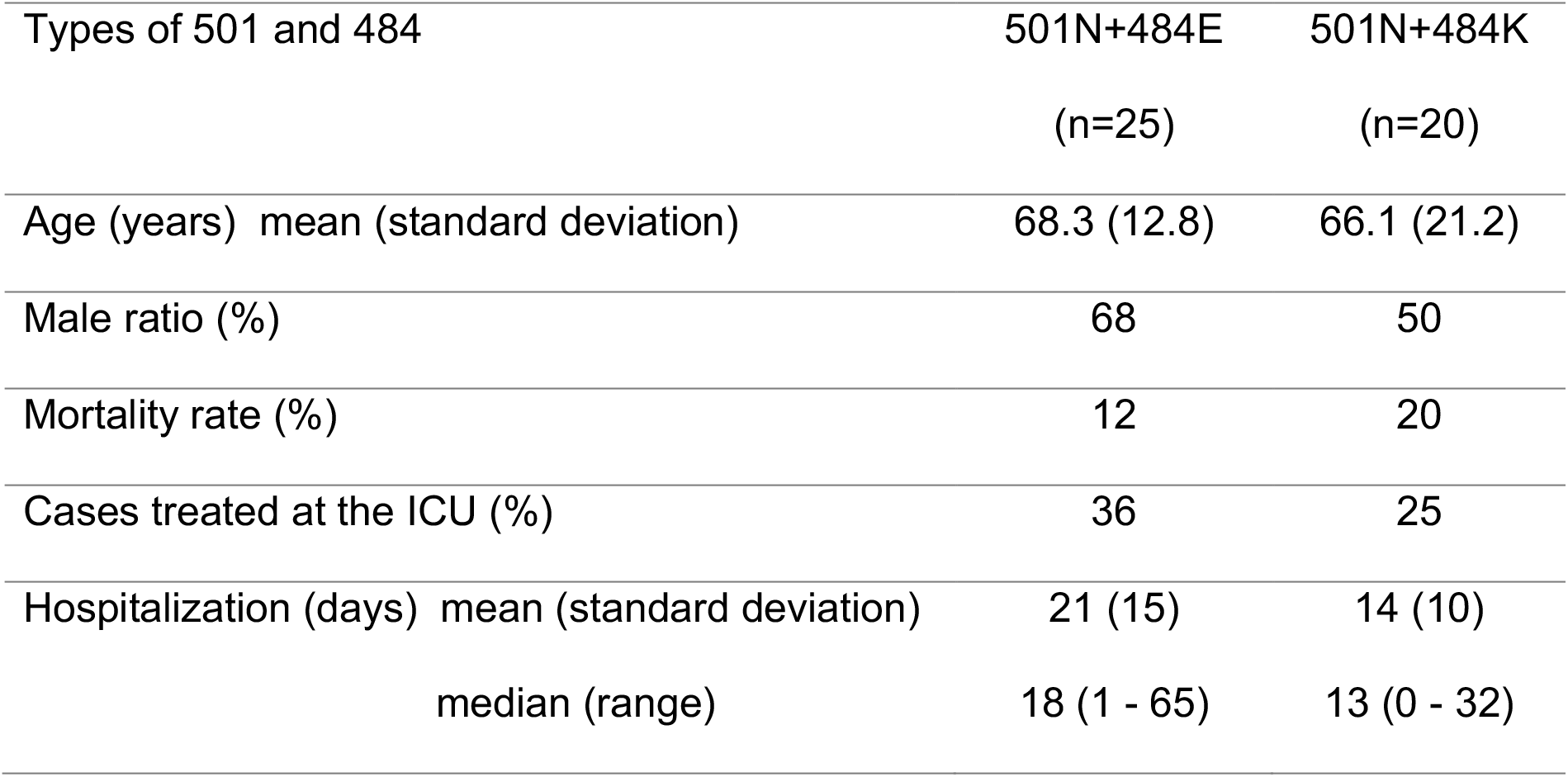
Clinical parameters of cases showing 501N+484E and 501N+484K types

## 4. Discussion

The B.1.1.7 variant is known to have high transmissibility and mortality.^1,2^ However, there are few reports about R.1 lineage. R.1 variants have also been detected in the United States and Europe according to the PANGO lineage database.^5^ However, the spread of R.1 variants during the pandemic in a particular area has not been reported.

In this study, we have found that the R.1 lineage variants rapidly prevailed in Tokyo in March 2021. The limitation of our study is that the subjects were limited to the patients who visited our hospital. However, we believe this finding also applies to Tokyo as a whole because the prevalence of R.1 variants in March was not only seen in our hospital, but also in a random sampling survey of patients in Tokyo recently performed by the Tokyo Metropolitan Institute of Public Health.^6^

Regarding the transmissibility of the R.1 variants, the fact that they rapidly replaced the B.1.1.214 variants (Japan lineage) in March suggests increased transmissibility. Because there were already many patients with COVID-19 in Tokyo in February, the founder effect of the R.1 variants seems unlikely. In a retrospective sampling analysis by WGS of 42 samples collected from November 2020 to January 2021, the R.1 variants were first detected in three samples collected in early January (data not shown).

Furthermore, R.1 variants were rapidly replaced by B.1.1.7 variants in April. This suggests that the B.1.1.7 variants have stronger infectivity, as previously reported.^1^ Interestingly, our study also revealed that there was no significant difference in copy numbers between the 501Y samples (UK lineage) and 501N samples (Japan lineage and R.1 lineage) as shown in Figure 1(B).

Regarding the severity of R.1 variants, there was no significant difference in mortality, ICU admission, and length of hospitalization between R.1 and B.1.1.214 cases, although the number of cases was small to be concluded. Our results suggest that the R.1 variants are not involved in the increased severity of the disease.

The E484K mutation has been reported to cause immune escape.^7^ The preventive effects of antibodies produced by vaccination can be diminished by E484K mutations. In fact, outbreaks of R.1 variants in a nursing facility that occurred after the residents and personnel were vaccinated were reported, although the infection rate of the vaccinated people was significantly lower than that of unvaccinated people.^8^

Since the vaccines are thought to be effective for the UK variants,^7^ it is possible that R.1 variants with immune escape might revive in the future. Just as the detection of the N501Y mutation and the survey of the B.1.1.7 variants are important, the detection of the E484K mutation and surveillance of the R.1 variants are still important for SARS-CoV-2 infection control.

## Data Availability

We will provide the PCR protocol and data on request.

## ACKNOWLEDGMENTS

We would like to thank all the staff involved in COVID-19 treatment that led to this study.

## AUTHOR CONTRIBUTIONS

Conceived and designed the study: KN, CTS, NI, YN, HT, ST. Analyzed samples: KN, CTS, YI, YT, SY, YT, JN, KS. Performed WGS: HT, KT, YT, AK. Analyzed data: NI, ST. Wrote the paper: ST.

